# Parental Dementia History is Associated with Peripheral Immune Aging Rather than Generalized Biological and Brain Age Acceleration in Midlife

**DOI:** 10.64898/2026.10.22.26366051

**Authors:** Edric D. Winford, Benjamin D. Huber, Justina F. Avila-Rieger, Calen P. Ryan, Mohamad J. Alshiko, Indira C. Turney, Jeffrey Pyne, Daniel W. Belsky, Adam M. Brickman, Jennifer J. Manly

## Abstract

Parental history of dementia is a risk factor for Alzheimer’s disease, although the mechanisms underlying this association remain poorly understood. We hypothesize that differences in epigenetic aging may contribute to this increased risk. We examined DNA methylation (DNAm)-based measures of epigenetic aging, telomere length, immune cell composition, and MRI-based measures of brain aging in 1,204 middle-aged adults. Parental dementia history was not associated with accelerated epigenetic or brain aging. However, it was associated with CD8 memory T cell expansion, with nominal differences in CD4 memory T cells, memory B cells, and telomere length. The association with shorter telomere length was attenuated after accounting for CD8 memory T cells, supporting a CD8-specific relationship. Earlier parental dementia onset was associated with faster pace of biological aging, while poorer parental cognitive performance was associated with lower naive and higher memory immune cell proportions, accelerated epigenetic aging, and a larger brain age gap. These findings suggest that midlife adults with parental dementia history exhibit a peripheral immune aging signature characterized by a naïve-to-memory T cell shift. Parental cognitive performance was also associated with accelerated epigenetic aging and brain age gap, suggesting that familial dementia risk is reflected across multiple aging dimensions in midlife offspring.

## Main

Years of age is the strongest risk factor for Alzheimer’s disease (AD), with incidence rates rising exponentially after age 65^1^. However, chronological age does not fully capture inter-individual variation in biological aging. Biological aging is reflected in epigenetic modifications, particularly DNA methylation (DNAm) patterns, which change with age and can be measured across multiple tissues to derive epigenetic clocks that serve as indicators of biological aging^2^. Epigenetic clocks measure various aspects of biological aging and were developed with different targets and training approaches. First-generation clocks, such as Horvath and Hannum, were trained on chronological age across multiple tissue types to predict biological age^3^. Second-generation clocks such as PhenoAge^4^ and GrimAge^5^ were trained on phenotypic variables and on plasma proteins and smoking pack years, respectively, to predict mortality risk and healthspan. Third-generation clocks, such as DunedinPACE^5^, do not estimate biological age but instead measure the rate at which an individual is aging over time; “pace of aging” epigenetic clocks are developed by modeling within-person longitudinal change in measurements of organ function. In AD, epigenetic clocks can predict progression from cognitively normal aging to mild cognitive impairment (MCI) or clinical AD^6,7^ and cognitive decline and incident dementia^8^. Furthermore, blood DNAm signatures can be detected years before clinical progression to dementia^9^. In human brain tissue, epigenetic age is associated with AD pathology and cognition^10^. These findings suggest that epigenetic modifications are associated with AD risk and pathophysiology, but heterogeneity across studies indicates that their role in clinical disease progression warrants further investigation^11,12^.

Brain age calculated with magnetic resonance imaging (MRI) captures age- and disease-related changes that differ from chronological age and is related to mortality risk. Typically, studies derive a brain age gap, defined as the difference between a participant’s predicted and chronological age, with a positive brain age gap indicating accelerated brain aging. Brain age is increased in individuals with MCI and can predict the progression from MCI to dementia^13^. DunedinPACNI was trained exactly the same as DunedinPACE, but using structural MRI data instead of DNAm^14^, and it predicts cognitive impairment, accelerated brain atrophy, and progression to dementia^15^. Together, these findings suggest that brain aging captures aspects of AD-related neurodegeneration that are detectable before clinical onset, making it a valuable complement to epigenetic measures for estimating biological aging in the context of AD and AD risk.

Individuals with a parent history of dementia have an increased risk of dementia and greater AD pathology^16^, poorer cognition,^17^ and lower microstructure in brain regions implicated in AD^18^compared with individuals without a parent history of dementia. Epigenetic aging profiles may therefore differ in individuals with a parental history of dementia and could represent a biological pathway through which intergenerational risk for AD is transmitted.

We examined biological aging in the Offspring study, a well-characterized, community-based cohort, to test the hypothesis that individuals with a parental history of dementia exhibit increased peripheral epigenetic and brain aging compared with those without a parental history of dementia. As DNA methylation-based deconvolution also enables estimation of peripheral immune cell composition, we also examined whether a history of parental dementia is associated with differences in immune cell populations. We previously showed that individuals with a parental history of dementia have increased pro-inflammatory cytokines^19^, and given that AD and AD risk are associated with changes in adaptive and innate immune response^20,21^, we hypothesized that immune cell composition may also differ in this population. We also examined whether parental age at dementia onset and parental cognitive performance are associated with indicators of biological aging.

## Results

We collected blood samples and derived DNAm data from 1,204 Offspring participants during their baseline study visit. Most participants identified as Hispanic (72%) or non-Hispanic Black (21%); 67% were female; and their ages ranged from 28 to 89 years (mean, 55.8 years). Compared with participants without parental history of dementia (N=776, 65%), those with a parental history of dementia (N=427, 35%) were older (mean age 59.6 vs 53.7), were more likely to be women (71% vs. 65%), to identify as Hispanic (87% vs. 63%), and to Speak Spanish as their primary language (70% vs. 36%), and had fewer years of education (mean 6.1 vs 5.9), higher rates of diabetes (24% vs. 17%), higher HbA1c (mean 6.1 vs. 5.9) and higher cardiovascular (CVD) burden (mean 0.8 vs. 0.6). There was no meaning full different in BMI between groups (mean 29.6 vs. 29.5). Of the 1,204 participants with DNAm data, 680 also had structural magnetic resonance imaging (MRI) data for calculating brain age indicators (**Supp Table 1**).

We first assessed whether individuals with a parental history of dementia exhibited accelerated epigenetic and brain aging compared to individuals without a parental history of dementia (**Figure 1**). After accounting for multiple testing with FDR correction, participants with parental dementia history did not differ from those without dementia history on any of the epigenetic clocks. We observed nominal differences in comparisons of DNAm-estimated telomere length (shorter in those with parental dementia history; β=-0.02, 95% CI [-.04, -.01], d = - 0.47) and GrimAge (younger in those with parental dementia history; β=-0.72, 95% CI [-1.3, -0.10], d=-0.16). GrimAge surrogate marker results are shown in **Supp Figure 1**. Results were similar in sensitivity analyses adjusting for CVD burden, a factor known to be associated with both dementia risk and epigenetic biomarkers, with the GrimAge association strengthening and the telomere length association attenuating after adjustment (**Supp Table 2**).

**Fig. 1.**
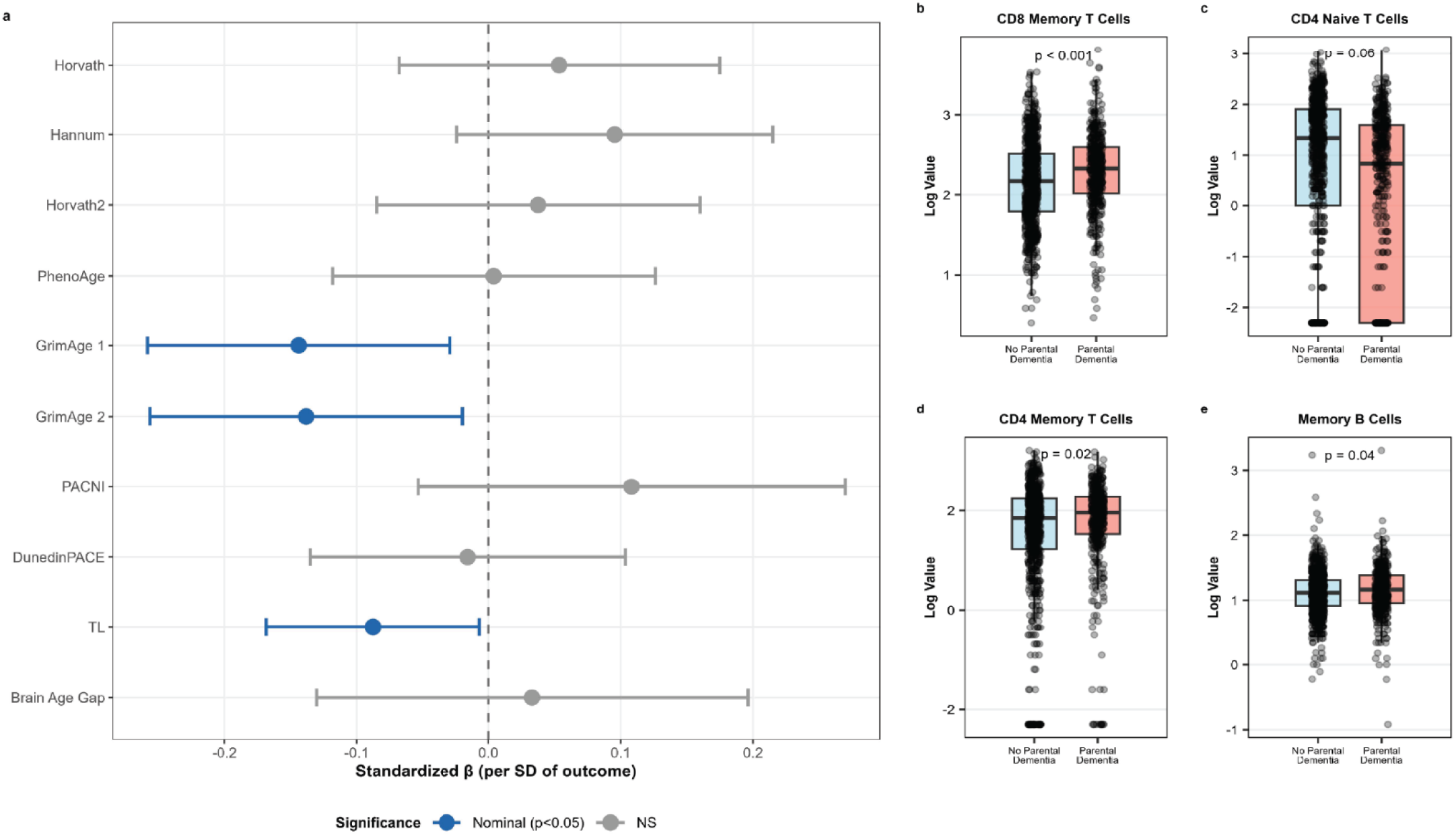
Epigenetic aging and peripheral immune cell composition in individuals with and without parental history of dementia. (a) Forest plot showing standardized regression coefficients (β per SD of outcome) for associations between parental dementia history and epigenetic clocks, telomere length, and brain aging measures, adjusted for age and sex. Error bars represent 95% confidence intervals. Blue indicates nominal significance (p<0.05), and gray indicates non-significant associations. (b–e) Boxplots comparing unadjusted log-transformed DNAm-estimated immune cell proportions between participants with no parental dementia history (blue) and those with parental dementia history (red) for (b) CD8 memory T cells, (c) CD4 naive T cells, (d) CD4 memory T cells, and (e) memory B cells. Boxes represent the interquartile range, with the median indicated by the horizontal line. P-values reflect linear regression models adjusted for age and sex.

Alterations in adaptive immunity are associated with AD and dementia risk^20,21^. DNAm-based aging measures are also partly driven by immune cell composition^22^. We therefore examined DNAm-estimated peripheral immune cell proportions and their relationship to parental dementia history and DNAm-estimated telomere length. We observed higher proportions of CD8 memory T cells (β=0.105, 95% CI [0.04, 0.17], d=0.28), CD4 memory T cells (β=0.16, 95% CI [0.02, 0.29], d=0.21), and memory B cells (β=0.04, 95% CI [0.01, 0.09], d=0.15), and lower proportions of CD4 naïve T cells (β=-0.18, 95% CI [-0.37, 0.01], d=-0.31) among individuals with a parental history of dementia compared to those without (Figure 3). After FDR correction, only CD8 memory T cells differed between groups (q=0.01) (**Figure 1 & Supp. Fig. 2**).

**Fig. 2.**
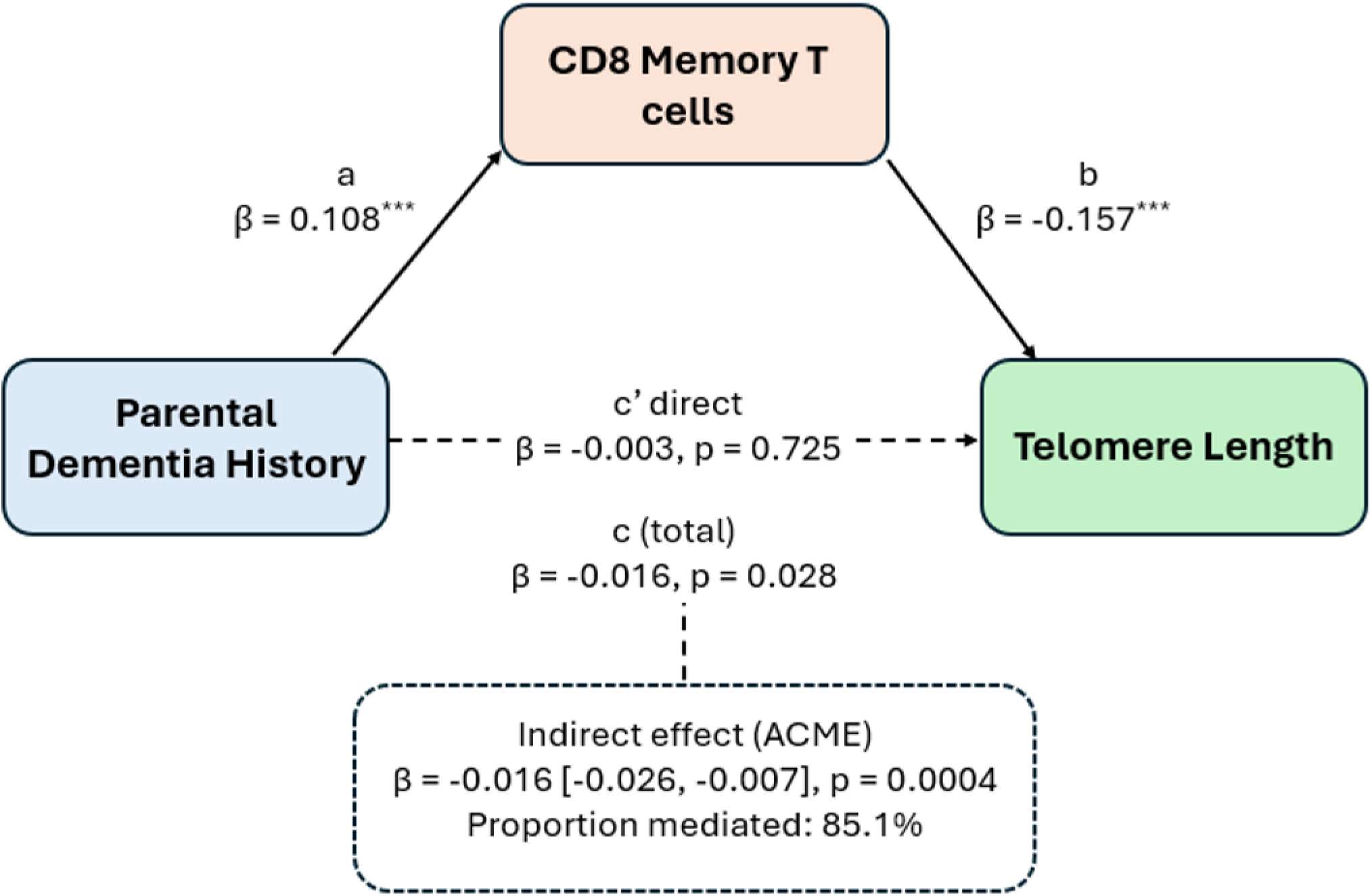
CD8 Memory T Cell Expansion Mediates the Association between Parental Dementia History and Telomere Attrition. Path diagram illustrating the mediation analysis examining CD8 memory T cell expansion as a mediator of the association between parental dementia history and telomere length (PCDNAmTL). Path a represents the association between parental dementia history and CD8 memory T cell expansion; path b represents the association between CD8 memory T cell expansion and telomere length; path c represents the total effect of parental dementia history on telomere length; and path c’ represents the direct effect after accounting for CD8 memory T cell expansion. The indirect effect (ACME) reflects the portion of the total effect operating through CD8 memory T cell expansion. All models were adjusted for offspring age and sex.

**Fig 3.**
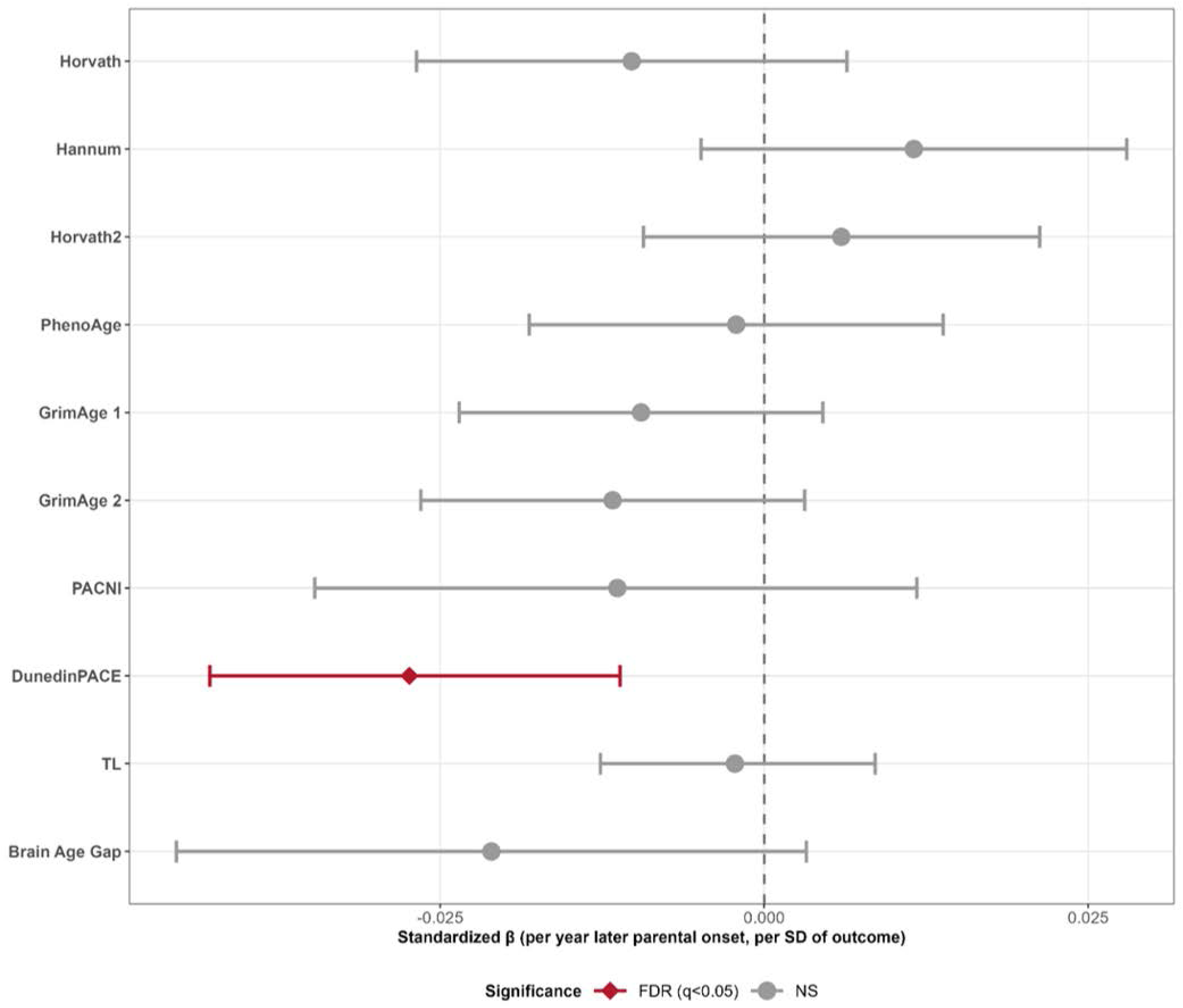
Associations with Parental Age of Dementia Onset and Epigenetic Clocks. Forest plot showing associations between parental age of dementia onset and offspring epigenetic clock measures, restricted to parental dementia-positive individuals only (n=331). Each point represents the standardized regression coefficient (β per year of later parental onset, expressed per standard deviation of each outcome) from models adjusted for offspring age and sex. Horizontal lines represent 95% confidence intervals. A negative beta indicates that later parental onset is associated with lower values of the outcome (i.e., slower biological aging). The red diamond indicates FDR-corrected significance (q<0.05); gray circles indicate non-significant associations.

DNAm-estimated immune cell composition drives variation in DNAm-based telomere length estimates^23^. We therefore examined whether CD8 memory T cell abundance accounted for the association between parental dementia history and DNAm-estimated telomere length. Mediation analyses indicated that CD8 memory T cell proportion accounted for 85.1% (95% CI [36.6%, 344.4%]) of the association between parental dementia history and DNAm-estimated telomere length (ACME = −0.017, 95% CI [−0.027, −0.007], p=0.0004), with no direct effect of parental dementia history on DNAm-estimated telomere length after accounting for CD8 memory T cell abundance (ADE = −0.003, 95% CI [−0.018, 0.012], p=0.725) (**Figure 2**). Consistent with these findings, adjusting for CD8 memory T cell proportion, but not CD4 memory T cell proportion, attenuated the association between parental dementia history and DNAm-estimated telomere length (**Supp Table 3**), supporting specificity to the CD8 memory T cell compartment.

To further characterize the relationship between parental dementia history and biological aging, we examined parental age at dementia onset and parental cognitive performance and their associations with biological aging outcomes and immune cell composition. Among individuals with a parental history of dementia, older parental age at dementia onset was associated with a slower pace of biological aging based on DunedinPACE (β=-0.003, 95% CI [-0.005, -0.001], d=-0.027) after FDR correction, but not with other epigenetic clocks (**Figure 3**). No associations were observed between parental age at dementia onset and immune cell composition (**Supp Figure 4**). As a sensitivity analysis, we examined whether parental age at onset, dichotomized at below and above the cohort median (age < 84.7 years), was associated with biological aging outcomes. Consistent with the continuous analysis, individuals with early parental onset had a faster pace of aging based on DunedinPACE compared with those with later parental onset (β=0.025, 95% CI [0.0002, 0.050]), which was nominally significant but did not survive FDR correction (**Supp Figures 5-7**). No other associations were observed with epigenetic clocks or immune cell composition.

**Fig. 4.**
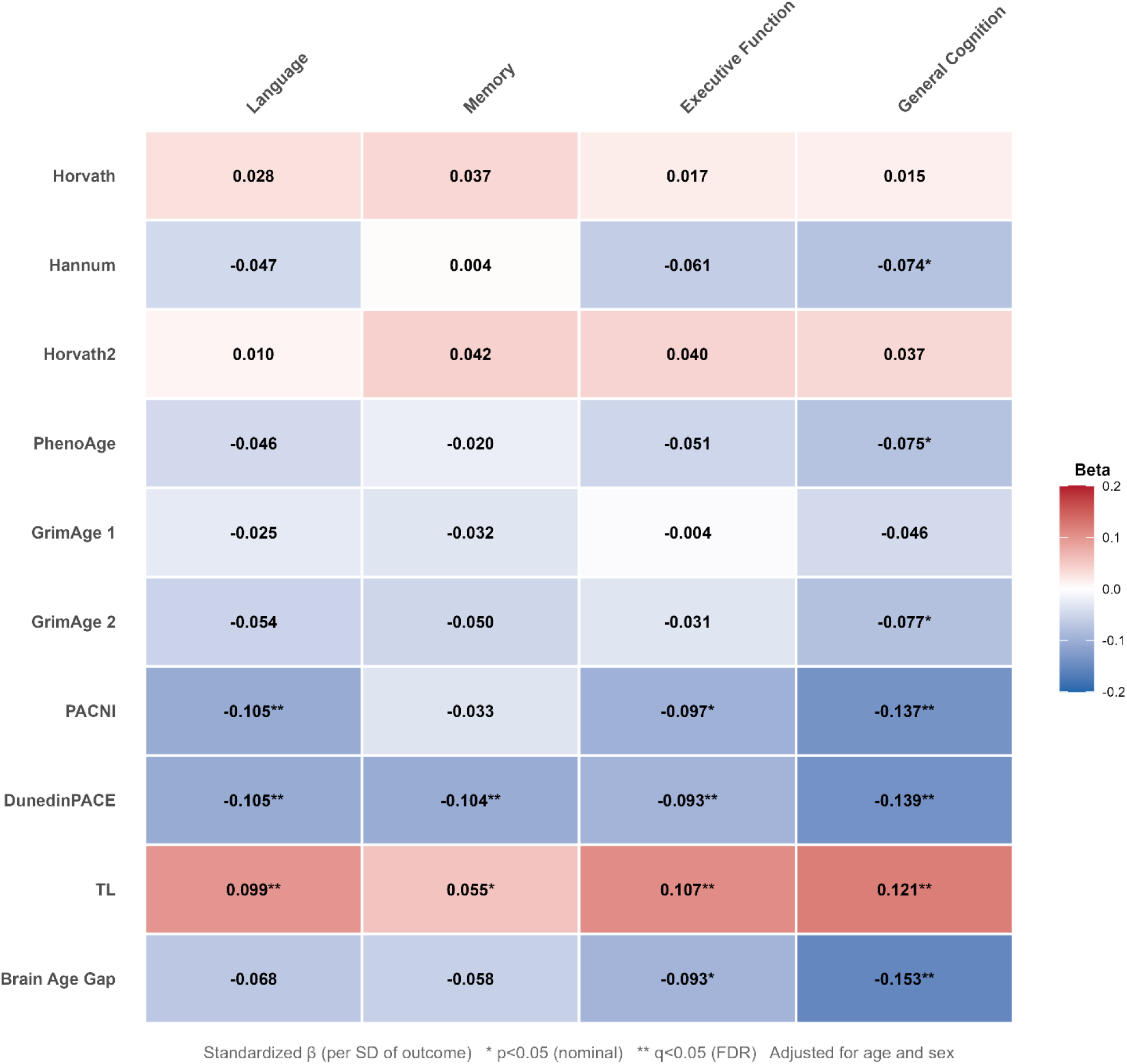
Associations between Parental Cognitive Factor Scores and Offspring Epigenetic Clock Measures. Heatmap showing regression coefficients (β) from models examining associations between parental cognitive factor scores (language, memory, executive function, and general cognition) and offspring epigenetic clock measures, adjusted for offspring age and sex. Each cell displays the standardized beta coefficient, with color intensity reflecting the magnitude and direction of the association; red indicates positive associations (higher parental cognition associated with higher clock values), and blue indicates negative associations (higher parental cognition associated with lower clock values). Asterisks denote significance level: *p<0.05 (nominal); **q<0.05 (FDR corrected).

**Fig. 5.**
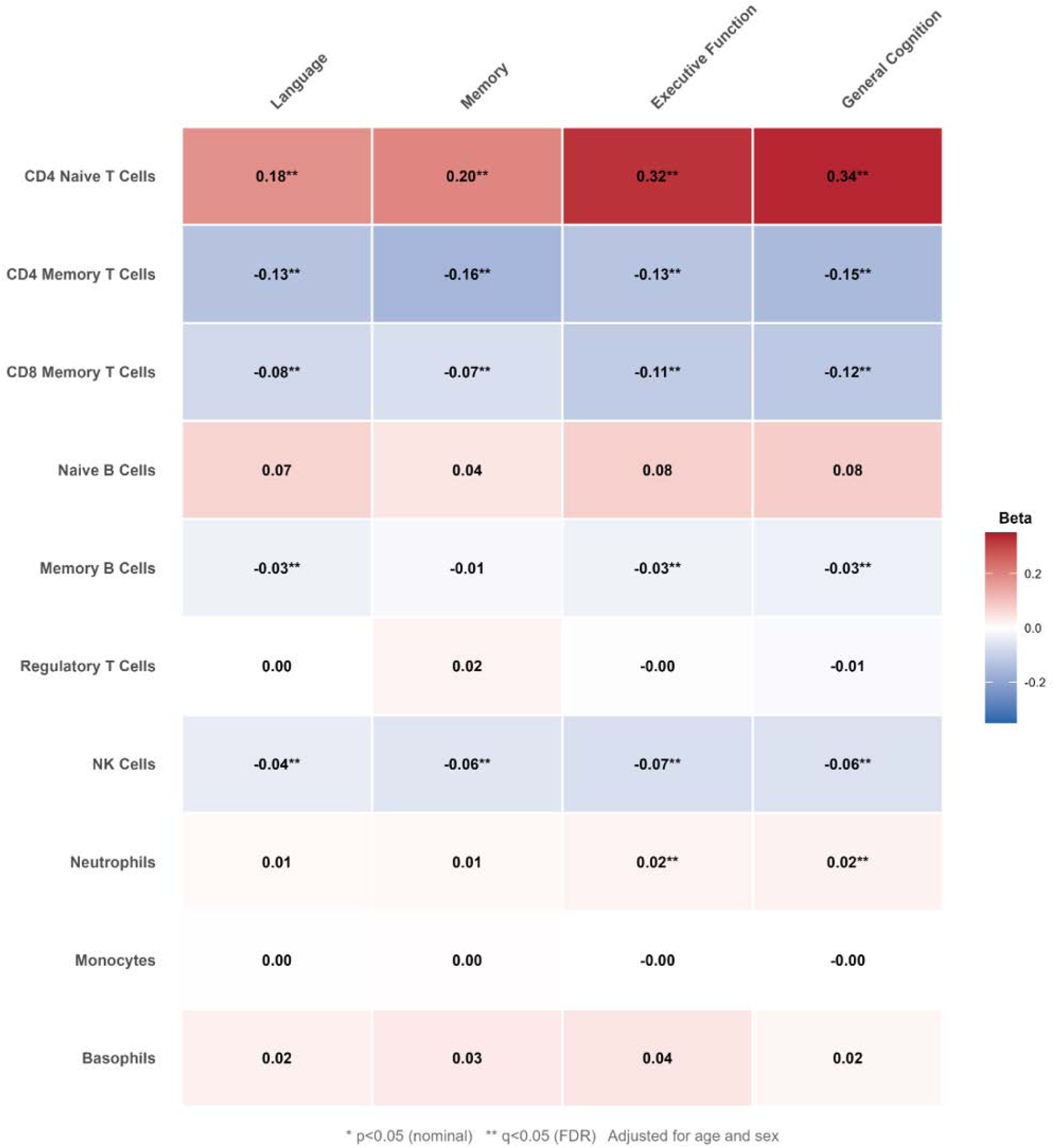
Associations between Parental Cognitive Factor Scores and Offspring DNAm-estimated Peripheral Immune Cell Composition. Heatmap showing standardized regression coefficients (β per standard deviation of outcome) from models examining associations between parental cognitive factor scores (language, memory, executive function, and general cognition) and offspring DNAm-estimated immune cell proportions, adjusted for offspring age and sex. Each cell displays the standardized beta coefficient, with color intensity reflecting the magnitude and direction of the association; red indicates positive associations (higher parental cognition associated with higher cell proportions), and blue indicates negative associations (higher parental cognition associated with lower cell proportions). Asterisks denote significance level: *p<0.05 (nominal); **q<0.05 (FDR corrected).

To extend beyond dementia status, we examined whether parental cognitive performance was associated with offspring biological aging. Higher overall parental cognitive performance, as captured by a general cognitive factor score derived from cognitive assessment measures, was associated with slower offspring pace of aging based on DunedinPACE (β=-0.139, 95% CI [-0.189, -0.089], d=-0.14) and DunedinPACNI (β=-0.137, 95% CI [-0.191, -0.083], d=-0.14), longer DNAm-estimated telomere length (β=0.121, 95% CI [0.071, 0.171], d=0.12), a lower brain age gap (β=-0.153, 95% CI [-0.207, -0.099], d=-0.15), lower Hannum acceleration (β=-0.074, 95% CI [-0.124, -0.024], d=-0.07), lower PhenoAge acceleration (β=-0.075, 95% CI [-0.125, -0.025], d=-0.08), and lower GrimAge 2 acceleration (β=-0.077, 95% CI [-0.127, -0.027], d=-0.08). After FDR correction, the associations with DunedinPACE, DunedinPACNI, DNAm-estimated telomere length, and brain age gap remained (all q<0.05) (**Figure 4**). With respect to GrimAge surrogate markers, results are shown in **Supplementary Figure 8**.

Parental cognitive performance was also associated with several DNAm-estimated peripheral immune cell populations. Offspring of parents with higher overall cognitive performance had lower CD4 memory T cell proportions (β=-0.15, 95% CI [-0.22, -0.08], d=-0.13), CD8 memory T cell proportions (β=-0.12, 95% CI [-0.15, -0.08], d=-0.23), memory B cell proportions (β=-0.03, 95% CI [-0.05, -0.01], d=-0.09), and NK cell proportions (β=-0.06, 95% CI [-0.10, -0.03], d=-0.11), as well as higher CD4 naive T cell proportions (β=0.34, 95% CI [0.23, 0.44], d=0.20), and neutrophil proportions (β=0.02, 95% CI [0.01, 0.03], d=0.10) (**Figure 5**).

## Discussion

We analyzed blood DNAm data to test whether middle-aged adults with a parental history of dementia showed signs of accelerated biological aging, differences in peripheral immune cell composition, and differences in MRI-based measures of brain aging relative to individuals without a parental history of dementia. After FDR correction, DNAm-estimated CD8 memory T cell proportions were higher among individuals with a parental history of dementia, and CD8 memory T cell abundance accounted for the association between parental history and DNAm-estimated telomere length. Among participants with a parental dementia history, earlier parental age at dementia onset was associated with a faster pace of aging as measured by DunedinPACE. In the full sample, lower parental cognitive performance was associated with older GrimAge, faster DunedinPACE, differences in DNAm-estimated immune cell composition, and MRI-based signs of accelerated brain aging.

We found that CD4 and CD8 memory T cells and memory B cells were higher in individuals with a parental history of dementia, with only CD8 memory T cells surviving FDR correction, whereas CD4 naïve T cells trended toward a decrease. Remodeling of the immune system is a hallmark of chronological aging, including shifts in T and B cell compartments from naïve to memory, as we observed nominally in our study^24–26^. However, as CD8 memory T cells were the only cell type robustly associated with parental history of dementia after FDR correction, these findings suggest a CD8-specific relationship with parental dementia history. Increased cytotoxic CD8^+^ T cells are positively associated with AD biomarkers and Aβ positivity, and negatively associated with cognitive decline^20,21,27^. They accumulate in the periphery in patients with AD^20,21^, and in individuals with mild cognitive impairment (MCI) and early Aβ accumulation, suggesting that their expansion precedes the onset of dementia. Most notably, CD8 T cells in dementia patients show increased expression of the senescence marker CD57, particularly within cytotoxic T cells; however, we are unable to delineate specific CD8^+^ senescent populations using DNA methylation. Nonetheless, these findings are consistent with a peripheral immune phenotype in individuals with a parental history of dementia that resembles observations reported in MCI and AD and is distinct from biological aging captured by epigenetic clocks. In our previous studies, we observed that individuals with a parental history of dementia had elevated pro-inflammatory cytokines, including Eotaxin-1 and IL-18, that increased with age, suggesting age-related alterations in immune function consistent with immune aging and cellular senescence; our current results showing DNAm-estimated shifts in immune cell composition provide additional evidence for this interpretation^19^. Notably, we previously showed that IL-18 levels increased with age, especially among individuals with a parental history of dementia, potentially reflecting accelerated inflammaging or a senescent profile, as senescent T cells express high levels of IL-18.

We next examined whether CD8 memory T cell abundance accounted for the association between parental dementia history and DNAm-estimated telomere length. The nominal association was largely accounted for by CD8 memory T cell abundance, consistent with the known dependence of blood-based telomere estimates on immune cell composition. Adjusting for CD8 memory T cell proportions fully attenuated the telomere length association, while adjusting for CD4 memory T cell proportions had minimal effect, supporting specificity to the CD8 memory compartment. As DNAm-estimated telomere length and immune cell proportions are derived from the same array, shared DNAm variance cannot be fully excluded. Nonetheless, if these findings reflect a true biological signal, CD8 memory expansion and DNAm-estimated telomere length are best understood as co-occurring features of an immune aging phenotype in individuals with a parental history of dementia, rather than reflecting a direct causal relationship. These findings suggest that peripheral immune aging, indexed by CD8 memory T cell expansion, may represent a biological signature of increased dementia risk in individuals with a parental history of dementia.

Whereas parental dementia status was most robustly associated with immune outcomes, parental cognitive performance was associated with a broader set of aging-related phenotypes in offspring, including a more naïve-skewed immune profile, longer DNAm-estimated telomere length, epigenetic pace of aging, and brain-age measures. Importantly, these associations extended beyond the CD8 memory T-cell findings observed for parental dementia status, suggesting that parental dementia diagnosis, age at dementia onset, and cognitive performance may capture distinct dimensions of familial vulnerability relevant to biological aging in offspring. Different parental cognitive and dementia-related characteristics provide complementary information about aging-related phenotypes in the next generation.

Across the epigenetic clocks, we found no differences in age acceleration with first-generation clocks (Horvath or Hannum) in individuals with a parental history of dementia; this observation is in line with previous studies showing that family history of AD is not associated with accelerated aging using these clocks^28^. We also did not find any differences in age acceleration for the second-generation clock PhenoAge or third-generation clock DunedinPACE, which measures the pace of aging. MRI measures of brain aging, such as brain age gap and DunedinPACNI, also showed no differences. Overall, these data suggest that parental history of dementia is not associated with accelerated brain or biological aging, as measured by these clocks, in middle-aged adults. In contrast, parental history of dementia was nominally associated with shorter DNAm-estimated leukocyte telomere length. Telomere shortening is associated with cellular senescence^29–31^. Shorter telomere length is associated with higher mortality rates, a greater risk of age-related diseases, and is causally linked to increased cognitive impairment, AD, and brain structural changes^32–38^. Furthermore, in middle-aged adults, shorter telomere length is associated with an increased risk of dementia^39^. As discussed, this association was largely accounted for by CD8 memory T cell abundance, consistent with the known relationship between immune cell composition and blood-based telomere estimates.

Several limitations should be noted. Although parental dementia history provides a clinically meaningful indicator of familial susceptibility to dementia, it does not allow us to determine the specific mechanisms through which risk is transmitted across generations; the biological signatures observed here may reflect a combination of shared genetic factors, environmental exposures, lifestyle characteristics, and aging-related processes that collectively shape dementia risk. Our sample is based in New York City, which may limit generalizability to other populations. Although we observe differences in DNAm-estimated telomere length, DNA methylation-based telomere estimation has a weak correlation with PCR-based measurement. However, DNAmTL is more strongly associated with age and mortality than qPCR-based leukocyte telomere length, suggesting it captures biologically meaningful signal despite this discordance^40^. Additionally, immune cell subset data derived from DNA methylation-based deconvolution should be considered exploratory, as these are estimated proportions rather than actual flow cytometry counts, which would be needed to verify immune populations. Finally, the proportion mediated by the CD8 memory T cell mediation analysis should be interpreted with caution, as the wide confidence interval reflects statistical instability when the total effect is small.

Overall, in a racially and ethnically diverse, well-characterized cohort with prospectively documented parental dementia status, our findings provide evidence that middle-aged individuals with a parental history of dementia exhibit a peripheral immune-aging signature that is distinct from generalized biological or brain aging. Parental dementia history was most robustly associated with CD8 memory T-cell expansion and was also nominally associated with shorter DNAm-estimated telomere length. The strong attenuation of the telomere association after accounting for CD8 memory T cell abundance suggests that these measures may reflect related aspects of peripheral immune aging rather than independent processes. Importantly, associations differed depending on how parental dementia risk was characterized, with three complementary exposures each capturing a different resolution of risk. Earlier parental dementia onset was associated with a faster pace of aging in offspring, while poorer parental cognitive performance was associated with a more memory-skewed immune profile, shorter telomere length, faster biological aging, and greater brain aging. Together, these findings suggest that parental dementia risk is reflected across multiple dimensions of aging in offspring and that different operationalizations of parental risk capture complementary aspects of immune, biological, and brain aging in midlife.

## Methods

### Participants

The Offspring Study is a community-based longitudinal study of aging and cognition aimed at identifying social, structural, and biological factors that contribute to racial and ethnic disparities in AD. Individuals enrolled in the Offspring Study are the adult children of participants in the Washington Heights Inwood Columbia Aging (WHICAP), a cohort of eligible adults 65 years or older followed longitudinally, for whom AD status is well characterized. Eligible Offspring participants are at least 28 years old, fluent in English or Spanish, and consent to provide blood samples. Participant evaluations occurred between January 2017 and March 2020. All participants completed informed consent forms, and the Columbia University Institutional Review Board approved all study protocols.

### Parental Dementia Status

Parental dementia status was determined through diagnostic consensus conferences conducted as part of WHICAP, as previously described^19,41,42^. Briefly, clinical diagnoses of dementia were assigned according to standard research criteria^43^, based on cognitive testing, a medical and neurological exam, and functional assessments, by a consensus panel including at least one neurologist and one neuropsychologist, blinded to previous diagnoses. Only incident dementia cases were included in our study; parents who were cognitively unimpaired at baseline and subsequently received a dementia diagnosis during follow-up. Parental age of dementia onset was defined as the age at dementia incidence visit.

### DNA Data Collection

Blood samples were collected into Becton Dickinson (BD) Vacutainer tubes containing ethylenediaminetetraacetic acid (EDTA). Samples were centrifuged at 2000 × g for 15 minutes at room temperature. The resulting plasma was aliquoted into polypropylene cryotubes and stored at −80 °C until analysis.

### DNA Methylation Processing and Epigenetic Clock Calculation

Preprocessing and normalization of DNA methylation data were conducted in R (v 4.4.1) on n=1248 samples. Samples with the following criteria were removed: samples methylated or unmethylated signal intensity <10.5 (n=11); samples with >5% of probes with fewer than 5 beads (n=2); samples with bisulfite conversion efficiency <80% (n=11), samples with an average detection p-value ≥0.05; samples with irregular clustering of SNP-based probes (minfi v1.5.0)^44^. Additionally, 17 metrics implemented in the ewastools package (v 1.7.2)^45^ were applied, with one sample (n=1) failing on non-polymorphic green channel intensity. An n=26 sample was removed due to a batch effect significantly impacting beta value distributions for those samples.

Normalization was performed using the preprocessNoob function in the minfi package (v1.50.0). Epigenetic clocks were calculated on Noob-normalized data using the methylClock package (v1.10.0) for original clocks and using methods described by Higgins-Chen et al^46^. DunedinPACE was derived as per Belsky et al^47^. Cell composition was estimated via a pipeline customized for EPICv2 arrays, and using the minfi estimateCellCounts2 function and the Salas extended reference panel^48^. Technical principal components were based on principal components analysis of control probes from samples that passed QC ^49^.

Probe QC was carried out on 930,075 probes. Probes were removed if they contained SNPs at the CpG site or single-base extension, were flagged as cross-hybridizing, or polymorphic^50,51^, mapped to X or Y chromosomes, had beadcounts <3 in >5% of samples (WateRmelon v2.10.0), or showed an average detection p-value ≥0.05 in >1% of samples. After QC, 862,599 probes remained for analysis.

### Brain age-gap prediction

Brain-predicted age was estimated using brainageR v2.1 (https://github.com/james-cole/brainageR), an open-access tool for generating brain-predicted age from raw T1-weighted MRI scans^52–54^. T1-weighted MRI scans from 1,064 Offspring participants were acquired on a 3T Siemens Prisma MRI scanner at the Zuckerman Mind Brain Behavior Institute and processed through the brainageR pipeline, which involves two main stages: pre-processing and prediction. In the pre-processing stage, images were segmented and spatially normalized using SPM12. Normalized gray matter, white matter, and cerebrospinal fluid maps were masked using a 0.3 threshold from the mean image template derived from the brainageR training dataset, then vectorized and combined. In the prediction stage, brainageR applies principal component analysis, retaining components capturing 80% of variance (435 components), and uses the resulting rotation matrix to project new data into the trained feature space. Brain-predicted age is then estimated using Gaussian process regression, trained on 3,377 healthy individuals (age range 18–92 years) from seven publicly available datasets. For each participant, brainageR outputs a brain-predicted age value. Brain age gap was calculated as the difference between predicted brain age and chronological age, with positive values indicating older predicted brain age relative to chronological age.

### Offspring Cardiovascular Burden

Participants were asked if they had ever been diagnosed with: Hypertension, Diabetes, Myocardial Infarction, Congestive Heart Disease, Other Heart Disease, and Stroke. Myocardial Infarction, Congestive Heart Disease and Other Heart Disease were then combined into a single variable indicating if they had been diagnosed with any of those conditions or none. We then added the number of cardiovascular conditions they had been diagnosed with, creating a cardiovascular burden score that ranged from 0 to 4.

### Parent Cognitive Performance

Parental cognitive performance was characterized by factor scores derived from neuropsychological testing conducted across WHICAP visits as previously described^41,42^. Briefly, three cognitive domains were assessed: memory, language, and executive function. Memory was assessed using the Selective Reminding Test (SRT), capturing total learning, delayed recall, and delayed recognition. Language was assessed using a 15-item version of the Boston Naming Test, and subtests of the Boston Diagnostic Aphasia Examination capturing repetition and comprehension. Executive function was assessed using the Similarities subtest of the Wechsler Adult Intelligence Scale-Revised, the Identities and Oddities subtest of the Mattis Dementia Rating Scale, and letter fluency. For each domain, individual test scores were standardized to z-scores using the sample mean and standard deviation at the initial occasion and averaged to create domain composite scores. A general cognition factor score was also computed by averaging across all three domain composites. Factor score means were calculated among WHICAP parents with three or more visits with a score for that domain who were cognitively unimpaired at the third visit. The mean of the first three eligible visits was used as the factor score, and parental age was calculated as the mean age across those three visits.

### Statistical Analysis

Associations between parental dementia status and DNAm-derived measures of epigenetic aging, telomere length, immune cell composition, and GrimAge surrogate biomarkers were examined using linear regression models adjusted for offspring age and sex. To examine whether clocks reflected intrinsic cellular or immune cell composition differences, all epigenetic clock models were re-adjusted for estimated immune cell proportions. CD8 naive T cells and eosinophils estimated from DNAm were excluded from all analyses due to over 80% of values having a zero value, rendering these estimates biologically unreliable. Parental dementia status was modeled as a binary exposure. Unstandardized regression coefficients (β), 95% confidence intervals, and p-values are reported for all primary analyses. Standardized effect sizes are reported as Cohen’s d for binary predictors and standardized beta coefficients for continuous predictors. Multiple comparisons were addressed using the Benjamini-Hochberg false discovery rate (FDR) procedure, with q < 0.05 considered statistically significant. Nominally significant associations (p < 0.05) are also reported where relevant. Sensitivity analyses were conducted to examine whether associations between parental history of dementia and DNAm outcomes were robust to adjustment for offspring CVD burden; models were re-run with CVD burden as an additional covariate, and changes in the primary dementia status coefficient are reported. The association between parental age of dementia onset and offspring biological aging outcomes was examined in the dementia-positive subsample only (n = 331), using continuous parental age of onset as the exposure in linear regression models adjusted for offspring age and sex. A sensitivity analysis using a median split (early onset: age < 84.7 years; late onset: age ≥ 84.7 years) was conducted to examine whether relative onset timing within the cohort was associated with biological aging. Associations between parental cognitive factor scores and offspring biological aging outcomes were examined using linear regression models adjusted for offspring age and sex in the full analytic sample. Mediation analysis was conducted to examine whether CD8 memory T cell expansion mediated the association between parental dementia history and telomere length, using the R package *mediation* with 1,000 bootstrapped iterations. The average causal mediation effect (ACME), average direct effect (ADE), total effect, and proportion mediated are reported with 95% bootstrapped confidence intervals. To further assess whether the association between parental dementia history depended on CD8 memory T cell abundance specifically, we additionally adjusted the telomere length model for CD8 memory T cell proportions and separately for CD4 memory T cell proportions as a specificity control. All statistical analyses were conducted in R.

## Data Availability

Data from this study are available upon reasonable request to the corresponding author, subject to review and approval by the investigative team.

## Acknowledgement

We would like to acknowledge the Offspring Study participants, the WHICAP participants, and the Offspring Study and WHICAP teams for their contributions to this study.

## Consent Statement

All participants in the Offspring and WHICAP study provided written informed consent.

