## Supplementary File for "Parental Dementia History is Associated with Peripheral Immune Aging Rather than Generalized Biological and Brain Age Acceleration in Midlife"

| **Supp Table 1. Baseline characteristics of the study sample** | | | |
| --- | --- | --- | --- |
| ***Offspring Sample*** | **Total** | **No Parental dementia** | **Parental dementia** |
| **Characteristic** | **(N=1204)** | **(N=776)** | **(N=427)** |
| Age, mean (SD) | 55.8 (11.3), 28.0-89.0 | 53.7 (11.0), 28.0-88.0 | 59.6 (10.8), 29.0-89.0 |
| Sex/Gender, N (%) |  |  |  |
| Female | 805 (67%) | 502 (65%) | 302 (71%) |
| Not Female | 398 (33%) | 273 (35%) | 125 (29%) |
| Race and Ethnicity, N (%) |  |  |  |
| Hispanic | 866 (72%) | 492 (63%) | 373 (87%) |
| Non-Hispanic Black | 244 (21%) | 198 (25%) | 46 (11%) |
| Non-Hispanic White | 87 (7.5%) | 79 (11%) | 8 (1.9%) |
| Other | 11(0.6%) | 7 (0.9%) | 4 (0.9%) |
| Primary Language, N (%) |  |  |  |
| English | 624 (52%) | 494 (64%) | 129 (30%) |
| Spanish | 580 (48%) | 282 (36%) | 298 (70%) |
| Years of Education, Mean (SD) | 12.8 (3.8), 0.0-20.0 | 13.4 (3.5), 0.0-20.0 | 11.6 (4.1), 0.0-20.0 |
| Unknown | 10 | 5 | 5 |
| Diabetes, yes N (%) | 223 (20%) | 125 (17%) | 98 (24%) |
| Hb1ac, Mean (SD) | 6.0 (1.3), 3.5-16 | 5.9 (1.2), 4.3-14 | 6.1 (1.4), 3.5-16 |
| BMI, Mean (SD) | 29.6 (6.6), 5.8-70.8 | 29.5 (6.8), 5.8-57.5 | 29.9 (6.2), 17.2-70.8 |
| CVD Burden, Mean (SD) | 0.6 (0.8), 0-4 | 0.6 (0.8), 0-4 | 0.8 (0.8), 0-3 |

**Supplementary Figures and Tables**

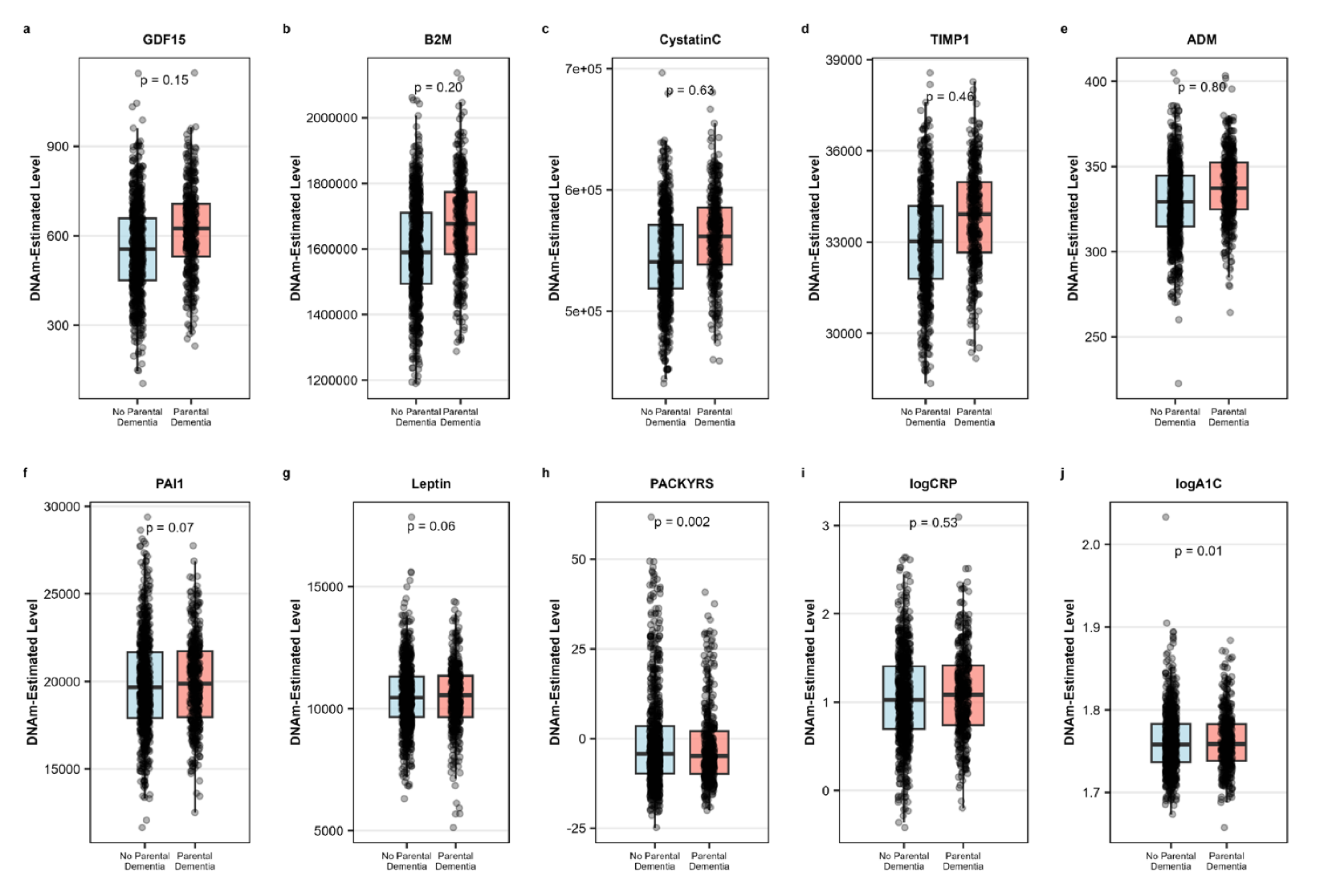

**Supp Fig. 1. GrimAge components in individuals with and without parental history of dementia.** Comparisons of unadjusted DNAm-based GrimAge component levels between participants with no parental dementia history (blue) and parental dementia history (red): (a) DNAmGDF15, (b) DNAmB2M, (c) DNAmCystatinC, (d) DNAmTIMP1, (e) DNAmADM, (f) DNAmPAI1, (g) DNAmLeptin, (h) DNAmPACKYRS, (i) DNAmlogCRP, and (j) DNAmlogA1C. Boxes represent the interquartile range with the median indicated by the horizontal lines. P-values reflect linear regression models adjusted for age and sex.

| **Supp Table 2.** Sensitivity analysis with CVD burden | | | | | |
| --- | --- | --- | --- | --- | --- |
| **Epigenetic clocks** | **Main Model β (95% CI)** | **p** | **CVD-Adjusted β (95% CI)** | **p** | **β Change** |
| Hannum | 0.42 [-0.11, 0.94] | 0.117 | 0.31 [-0.23, 0.86] | 0.259 | -0.10 |
| Horvath2 | 0.14 [-0.31, 0.58] | 0.548 | 0.17 [-0.29, 0.63] | 0.476 | +0.03 |
| PhenoAge | 0.02 [-0.64, 0.68] | 0.948 | -0.11 [-0.79, 0.57] | 0.752 | -0.13 |
| GrimAge 2 | **-0.72 [-1.34, -0.10]** | **0.022** | **-0.74 [-1.38, -0.11]** | **0.022** | **-0.02** |
| DunedinPACE | -0.002 [-0.016, 0.012] | 0.796 | -0.003 [-0.017, 0.011] | 0.633 | -0.002 |
| Telemore length | **-0.019 [-0.037, -0.002]** | **0.034** | -0.017 [-0.035, 0.002] | 0.084 | +0.003 |
| PACNI | 0.048 [-0.024, 0.119] | 0.190 | 0.045 [-0.028, 0.118] | 0.226 | -0.003 |
| **GrimAge Components** |  |  |  |  |  |
| GDF15 | -6.43 [-15.28, 2.42] | 0.154 | -6.29 [-15.48, 2.89] | 0.179 | +0.14 |
| B2M | 5023.9 [-2604.9, 12652.7] | 0.197 | 4477.0 [-3532.8, 12486.7] | 0.273 | -547.0 |
| CystatinC | 528.4 [-1625.5, 2682.2] | 0.630 | 678.9 [-1558.9, 2916.7] | 0.552 | +150.6 |
| TIMP1 | -29.8 [-108.0, 48.4] | 0.455 | -24.0 [-105.1, 57.2] | 0.562 | +5.8 |
| ADM | 0.23 [-1.52, 1.97] | 0.801 | 0.15 [-1.69, 1.98] | 0.874 | -0.08 |
| PAI1 | -285.8 [-597.4, 25.9] | 0.072 | **-322.7 [-630.8, -14.6]** | **0.040** | -37.0 |
| Leptin | -156.3 [-319.7, 7.0] | 0.061 | -120.2 [-286.9, 46.5] | 0.158 | +36.2 |
| PACKYRS | **-2.41 [-3.95, -0.88]** | **0.002** | **-2.42 [-4.00, -0.83]** | **0.003** | -0.001 |
| logCRP | -0.020 [-0.080, 0.041] | 0.527 | -0.022 [-0.085, 0.041] | 0.490 | -0.003 |
| logA1C | **-0.006 [-0.010, -0.001]** | **0.011** | **-0.006 [-0.010, -0.002]** | **0.003** | -0.001 |

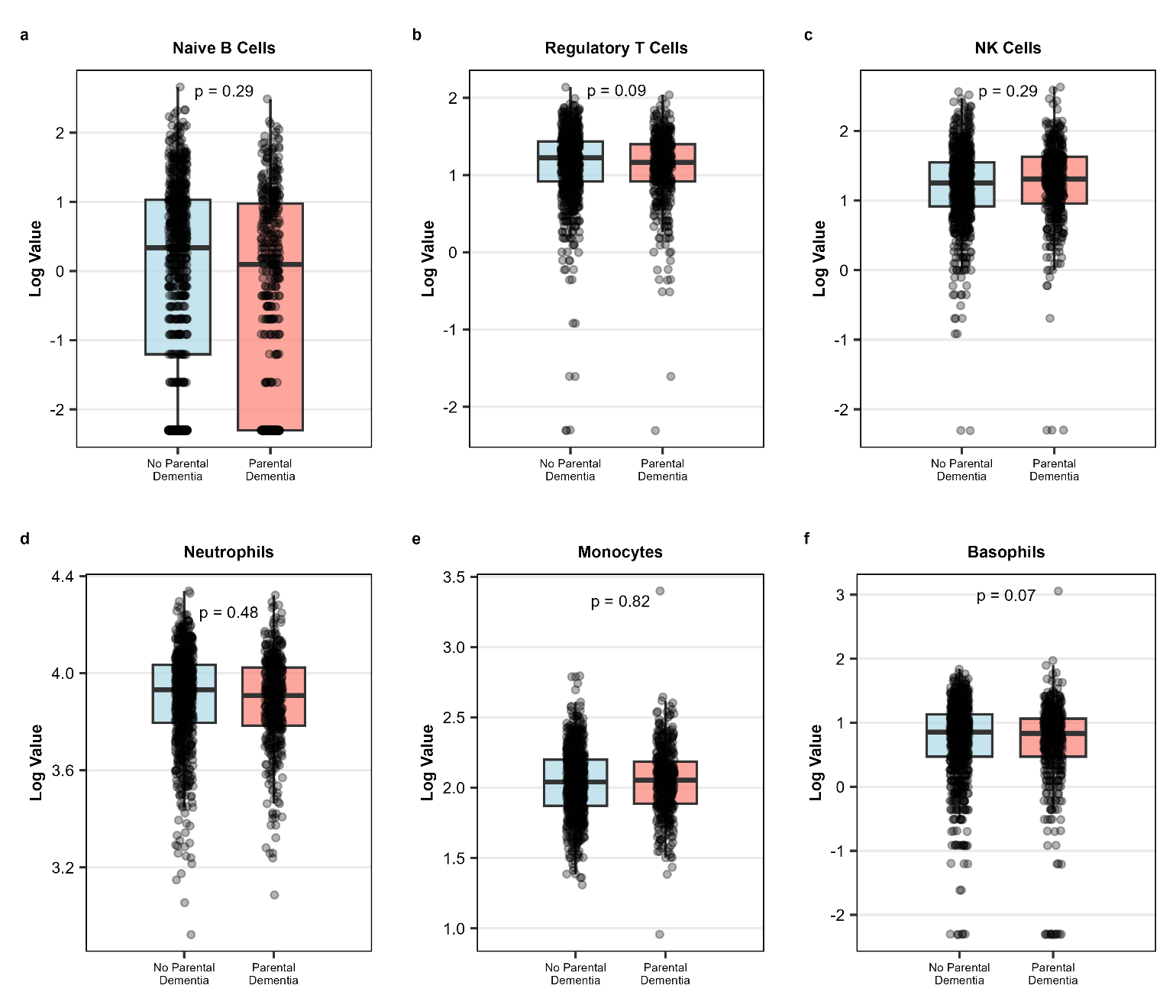

**Supp Fig. 2. DNAm-based peripheral immune cell composition in individuals with and without parental history of dementia.** Comparison of log-transformed DNAm-based immune cell counts between participants with no parental dementia history (blue) and parental dementia history (red) (a) naive B cells, (b) regulatory T cells, (c) NK cells, (d) neutrophils, (e) monocytes, and (f) basophils. Boxes represent the IQR with the median indicated by the horizontal line. P-values reflect linear regression models adjusted for age and sex.

| **Supp Table 3.** Sensitivity analysis examining whether the association between parental dementia history and telomere length depends on CD8 memory T cell abundance | | |
| --- | --- | --- |
| **Model** | **β (95% CI)** | **p-value** |
| TL ~ demstat + age + sex | -0.02 [-0.04, -0.002] | **0.034** |
| TL ~ demstat + age + sex + CD8 Memory T cells | -0.003 [-0.02, -0.012] | 0.707 |
| TL ~ demstat + age + sex + CD4 Memory T cells | -0.002 [-0.04, -0.001] | **0.044** |
| TL ~ demstat + age + sex + all immune cells | -0.001 [-0.02, 0.014] | 0.854 |

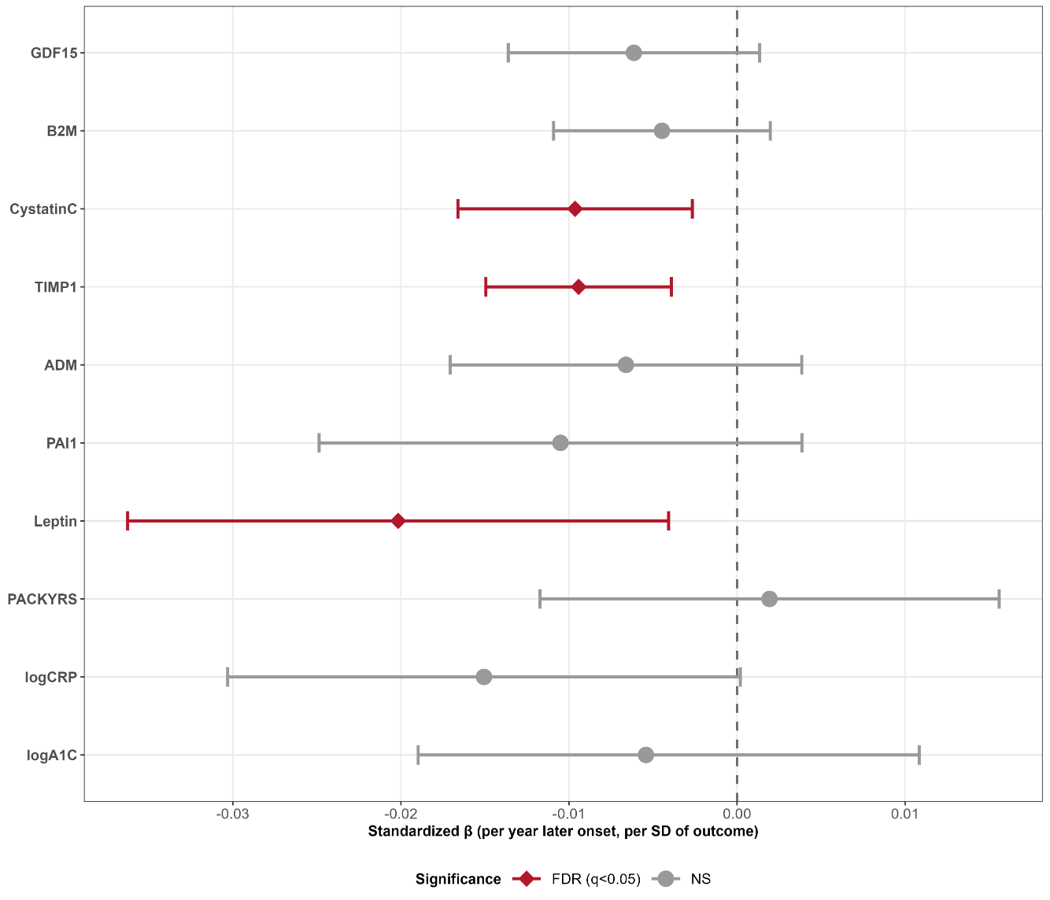

**Supp Fig. 3. Associations with Parental Age of Dementia Onset and GrimAge Surrogate Markers.** Forest plot showing associations between parental age of dementia onset and DNAm-estimated GrimAge surrogate markers in offspring, restricted to parental dementia-positive individuals only (n=331). Each point represents the standardized regression coefficient (β per year of later parental onset, expressed per standard deviation of each outcome) from models adjusted for offspring age and sex. Horizontal lines represent 95% confidence intervals. A negative beta indicates that later parental onset is associated with lower levels of the surrogate marker. The red diamond indicates FDR-corrected significance (q<0.05); gray circles indicate non-significant associations.

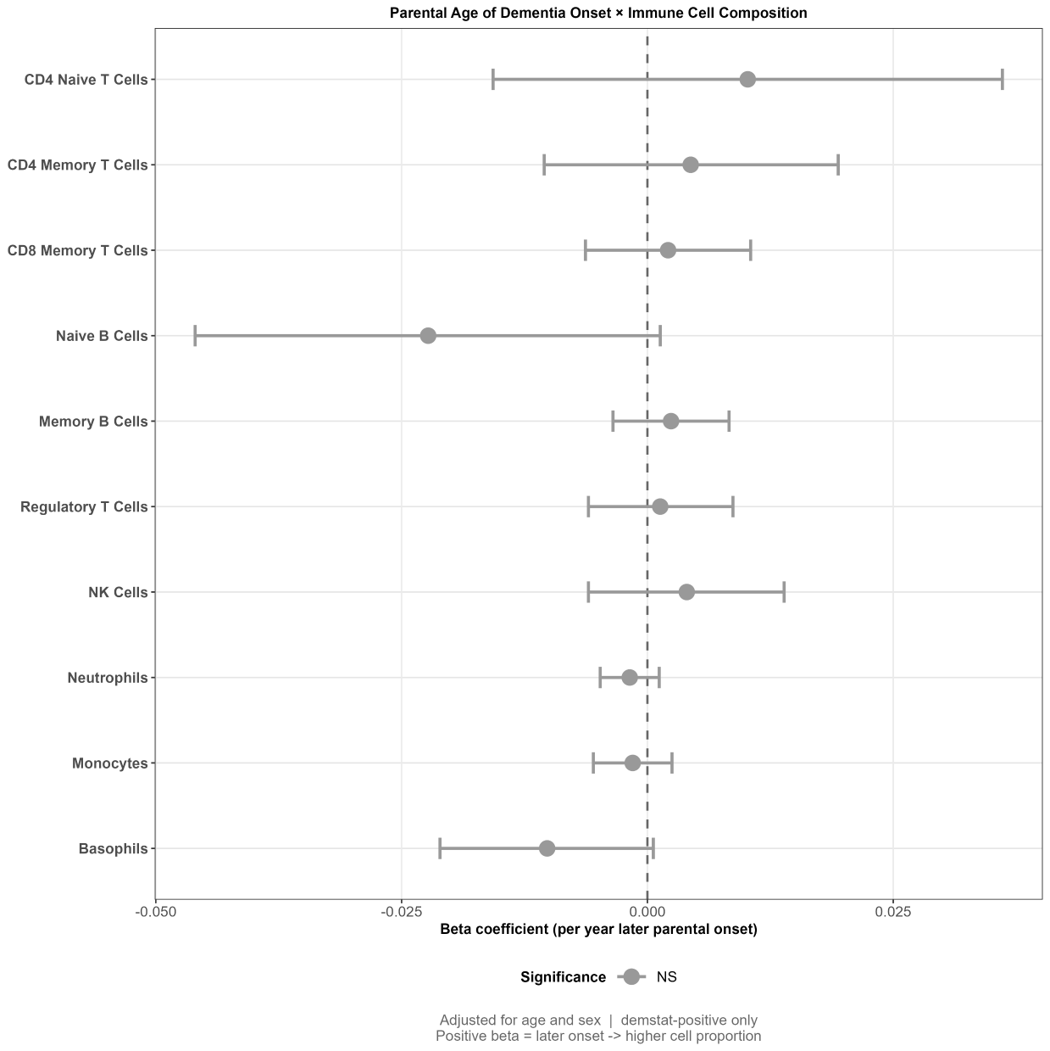

**Supp Fig. 4. Associations with Parental Age of Dementia Onset and Peripheral Immune Cell Composition.** Forest plot showing associations between parental age of dementia onset and DNAm-estimated immune cell proportions in offspring, restricted to dementia-positive individuals only (n=331). Each point represents the unstandardized regression coefficient (β per year of later parental onset) from models adjusted for offspring age and sex. Horizontal lines represent 95% confidence intervals. A positive beta indicates that later parental onset is associated with higher proportions of that immune cell population.

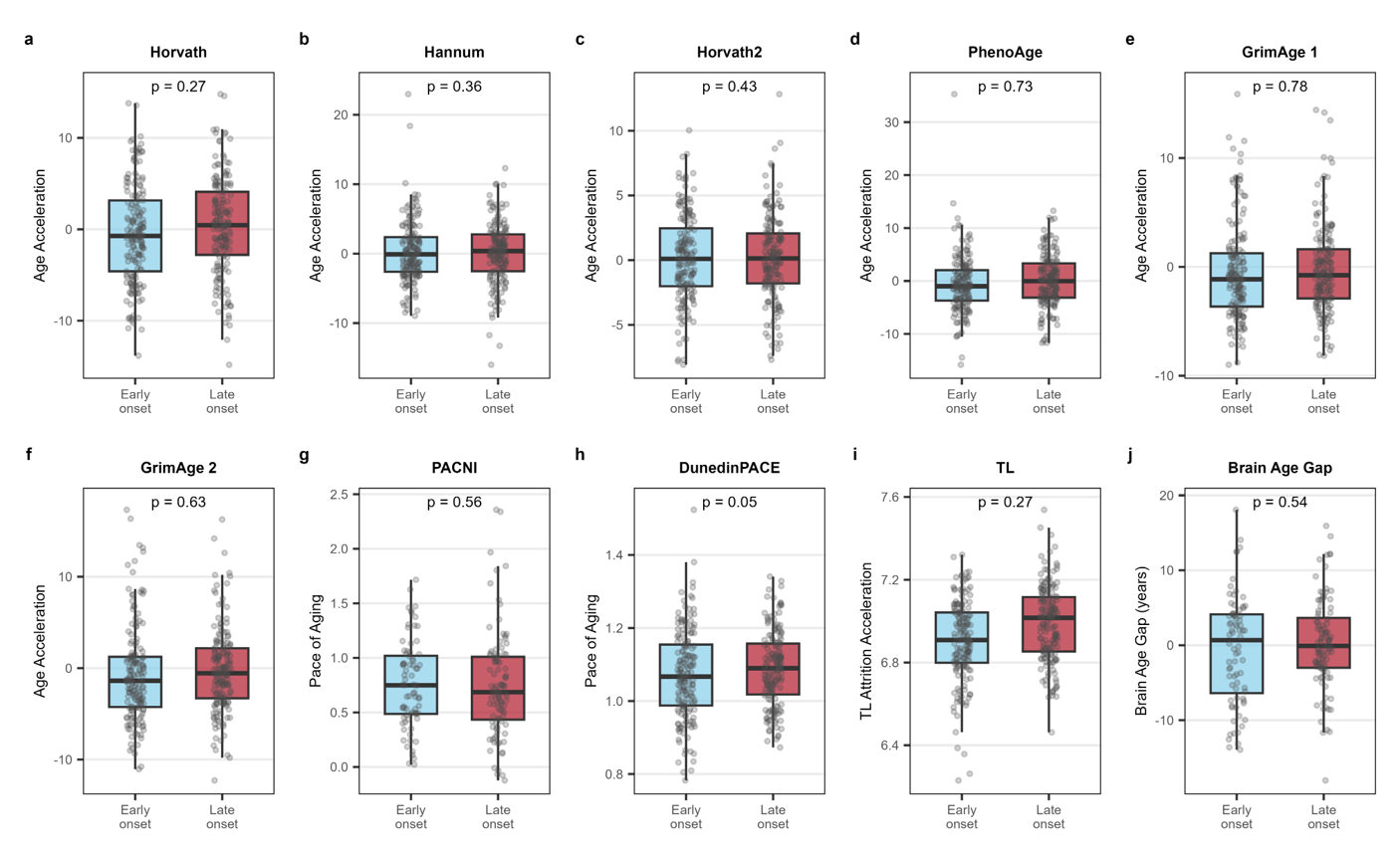

**Supp Fig. 5. Epigenetic Aging by Early versus Late Parental Age of Dementia Onset.** Boxplots comparing epigenetic clock measures between offspring of parents with early onset (below cohort median, age < 84.7 years; blue) and late onset (above cohort median, age ≥ 84.7 years; red) dementia, restricted to dementia-positive individuals only (n=331). Each panel displays the distribution of individual data points overlaid on the boxplot, with the median represented by the horizontal line and boxes representing the interquartile range. P-values reflect group differences from linear regression models adjusted for offspring age and sex. Early and late onset groups were defined relative to the cohort median parental age of dementia onset.

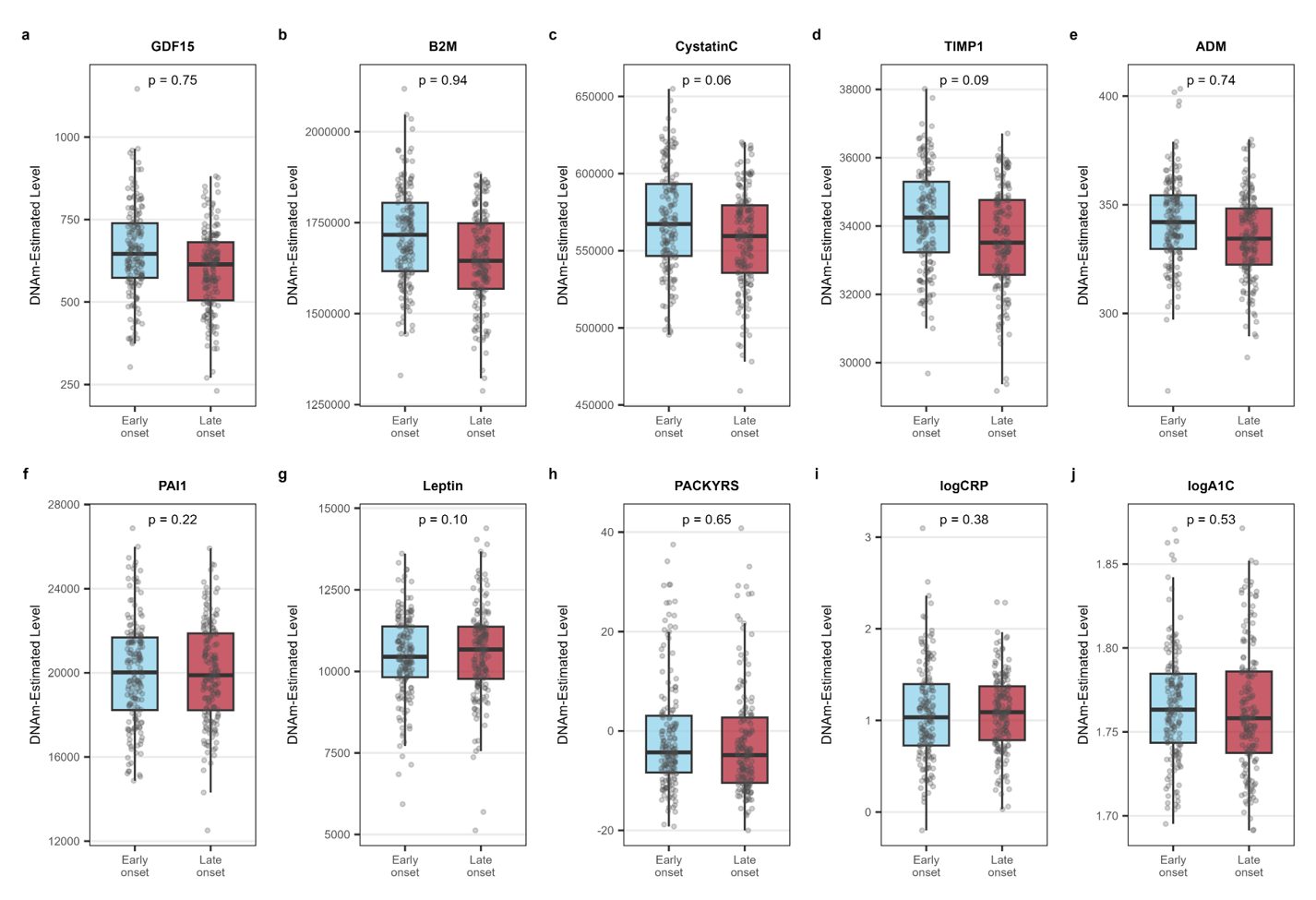

**Supp Fig. 6. GrimAge Surrogate Markers by Early versus Late Parental Age of Dementia Onset.** Boxplots comparing DNAm-estimated GrimAge surrogate marker levels between offspring of parents with early onset (below cohort median, age < 84.7 years; blue) and late onset (above cohort median, age ≥ 84.7 years; red) dementia, restricted to dementia-positive individuals only (n=331). Each panel displays the distribution of individual data points overlaid on the boxplot, with the median represented by the horizontal line and boxes representing the interquartile range. P-values reflect group differences from linear regression models adjusted for offspring age and sex. Early and late onset groups were defined relative to the cohort median parental age of dementia onset.

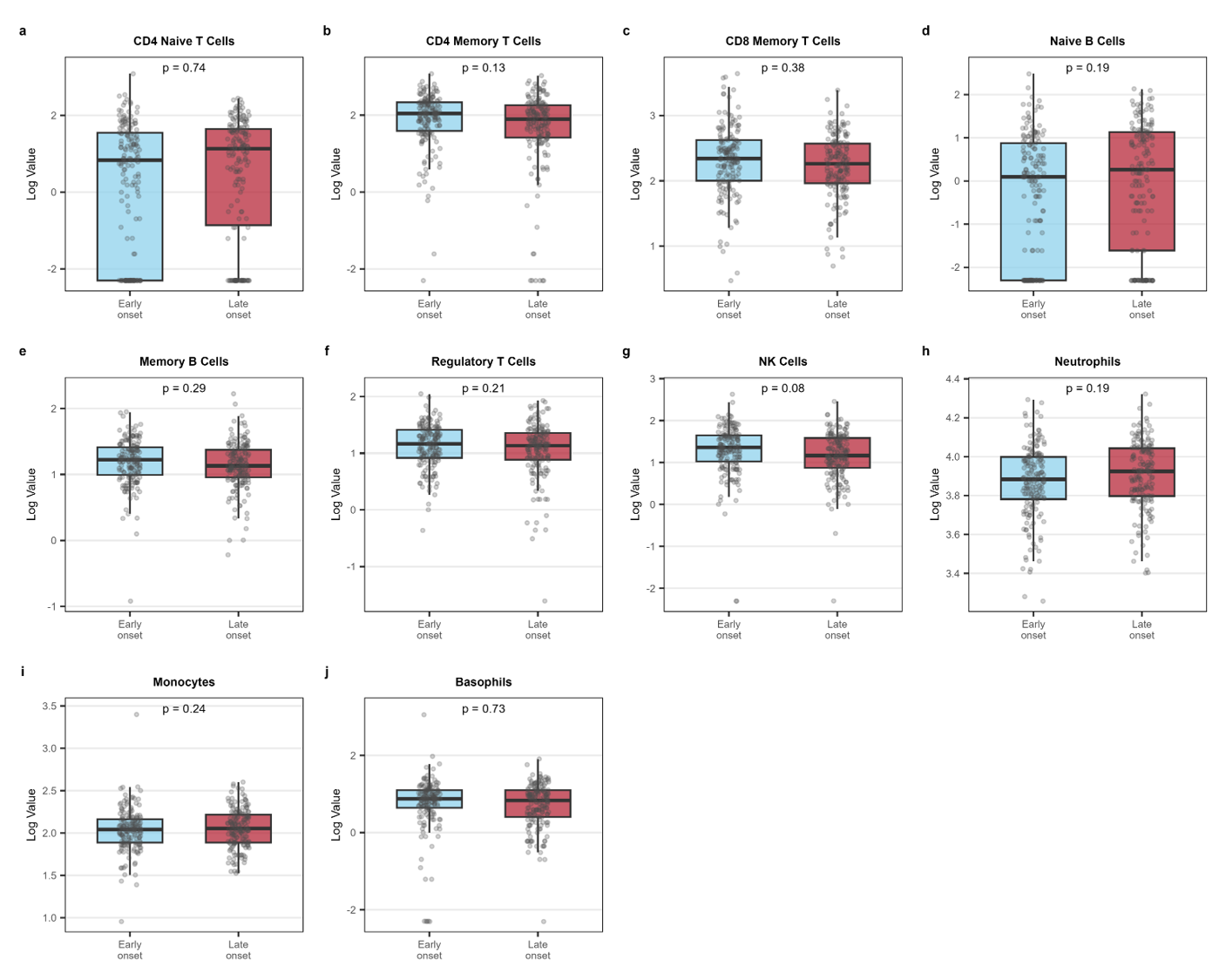

**Supp Fig. 7. Peripheral Immune Cell Composition by Early versus Late Parental Age of Dementia Onset.** Boxplots comparing DNAm-estimated immune cell proportions between offspring of parents with early onset (below cohort median, age < 84.7 years; blue) and late onset (above cohort median, age ≥ 84.7 years; red) dementia, restricted to dementia-positive individuals only (n=331). Each panel displays the distribution of individual data points overlaid on the boxplot, with the median represented by the horizontal line and boxes representing the interquartile range. P-values reflect group differences from linear regression models adjusted for offspring age and sex. Early and late onset groups were defined relative to the cohort median parental age of dementia onset.

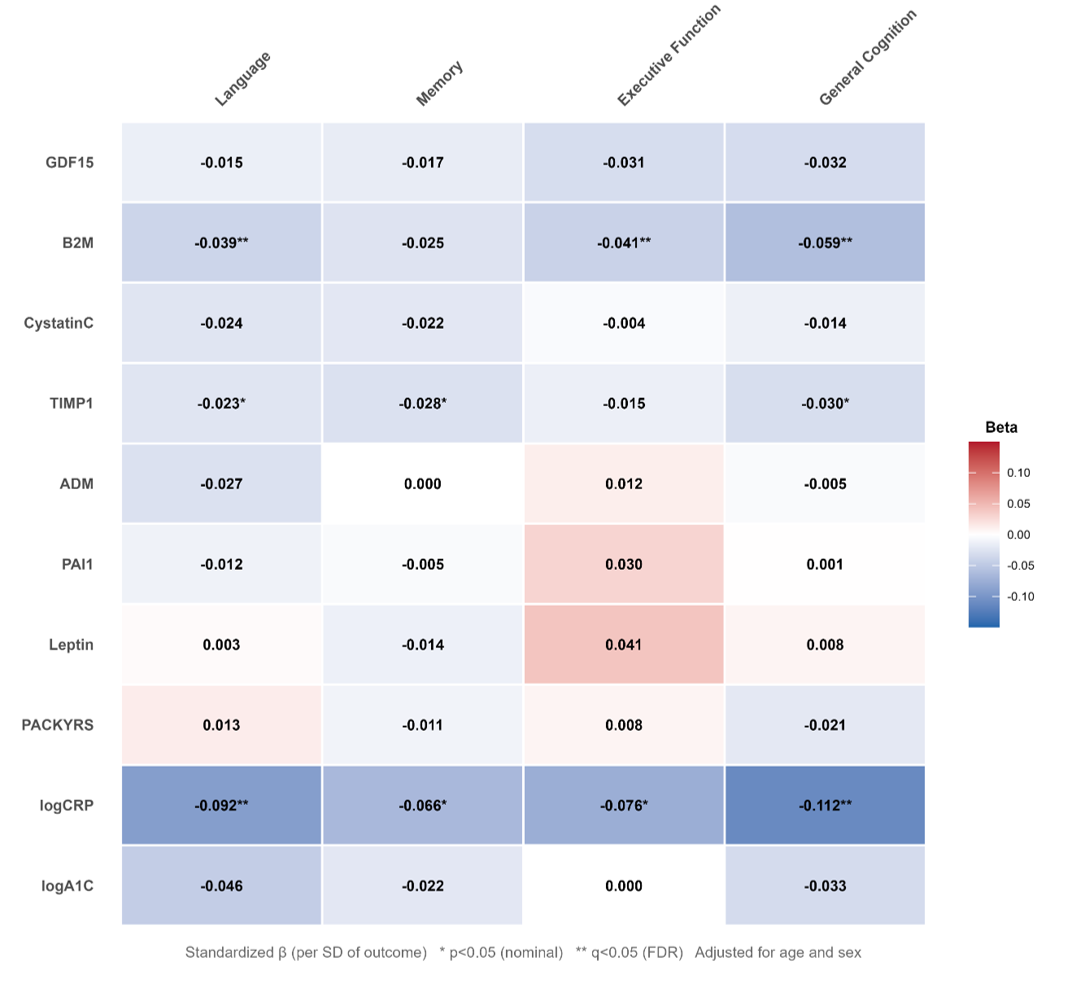

**Supp Fig. 8. Associations between Parental Cognitive Factor Scores and DNAm-Estimated GrimAge Surrogate Markers.** Heatmap showing standardized regression coefficients (β per standard deviation of outcome) from models examining associations between parental cognitive factor scores (language, memory, executive function, and general cognition) and offspring DNAm-estimated GrimAge surrogate markers, adjusted for offspring age and sex. Each cell displays the standardized beta coefficient, with color intensity reflecting the magnitude and direction of the association — red indicates positive associations and blue indicates negative associations (higher parental cognition associated with lower surrogate marker levels). Asterisks denote significance level: *p<0.05 (nominal); **q<0.05 (FDR corrected).
